# Logistic Formula in Biology and Its Application to COVID-19 in Japan

**DOI:** 10.1101/2021.03.24.21254279

**Authors:** Akira Kokado, Takesi Saito

## Abstract

A logistic formula in biology is applied, as the first principle, to analyze the second and third waves of COVID-19 in Japan.

## 1 Introduction

The logistic formula is useful in the population problem in biology. It is closely related to the SIR model [1] in the theory of infection [2–9]. In previous papers [10–13] we have shown that the logistic formula can be approximately driven from the SIR model. In the present paper, however, we regard the logistic formula as the first principle, which is independent of the SIR model. We follow the observation that the removed number *R*(*t*) in the SIR model behaves like the population in biology, that is, *R*(*t*) is a sum of accumulated numbers of deaths and discharged.

In Sec. 2 we give the logistic formula for *R*(*t*). Our policy is to use only data of removed by COVID-19 in Japan [14]. This formula will be applied to analyze the second and third waves in Secs. 3 and 4, respectively. These analyses revise results obtained in previous works [12,13]. The final section is devoted to concluding remarks. In Appendix we prepare error estimation formulas.

## 2 The logistic formula

In biology the logistic equation for a population *N* (*t*) is given by

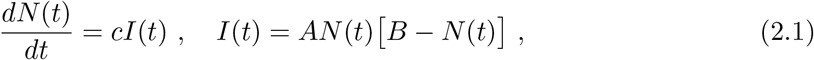

where *A, B* and *c* are some parameters. The solution is easily obtained as

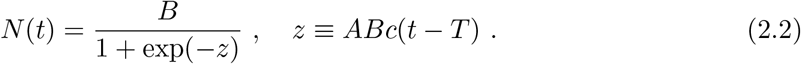

Here *T* gives a peak of *I*(*t*), which is given by

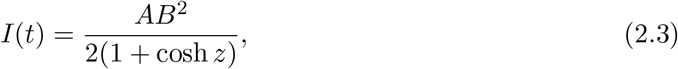

that is, *I*(*t*) = *AB*^2^*/*4.

The equation (2.1) can be regarded as the third equation of the SIR model,

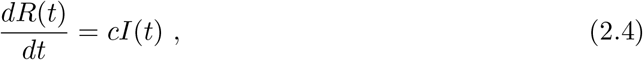

if we identify *N* (*t*) with *R*(*t*), where *R*(*t*) and *I*(*t*) are the removed number and the infectious number, respectively, and *c* the removed ratio. In previous works [10–13], our logistic formulas (2.2) and (2.3) have been driven approximately from the SIR model. In the present paper, however, we regard our logistic formulation as more fundamental rather than the SIR theory.

Let us rewrite Eq. (2.2) in notations *A* = *a/d, B* = *d* and *N* (*t*) = *R*(*t*) as follows:

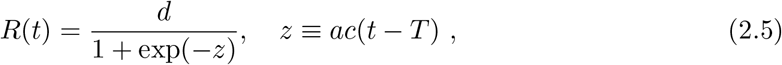

where *d* is the final total removed number, e. g., *d* = *R*(∞). Eq. (2.5) can be expressed as

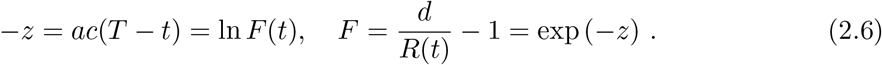

Accordingly, for different times *t*_*n*_, *t*_*n*+1_ and *t*_*n*+2_, (*n* = 1, 2, …), we have

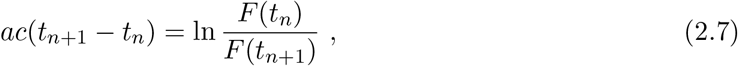

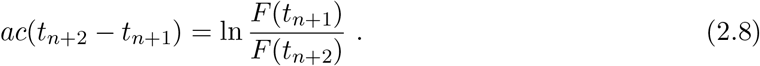

When time differences in Eqs. (2.7) and (2.8) are equal, we have a useful formula for *d*

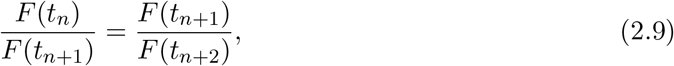

explicitly,

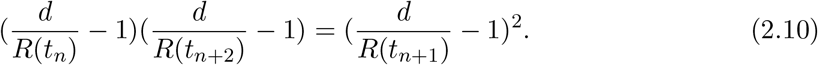

## 3 Application to the second wave of COVID-19 in Japan

Our logistic formula is applied to the second wave of COVID-19 in Japan. This provides a revise of previous work [12]

The *R*(*t*) is the accumulated number of removed in the second wave in Japan, which is an average for 7 days in a middle at each t with standard deviations, where t is the date starting from June 20, 2020. The virus is now called the Tokyo type. We have subtracted the accumulated number 20507 on July 19 of removed in the first wave from that in the first and second waves.

Substituting data in the Table 1 into Eq. (2.10) with *n* = 1, we have the equation for *d*

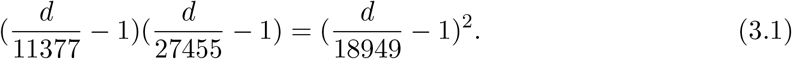

**Table 1:**
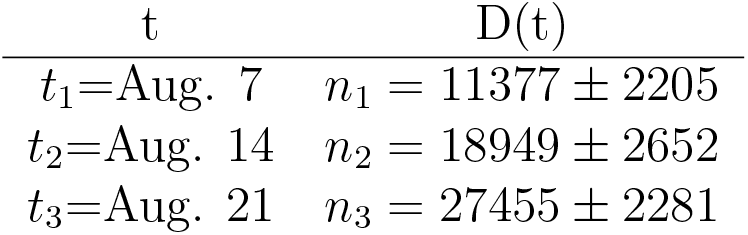
Date t and the removed number *R*(*t*) in the second wave in Japan [14]

| $t$ | $D(t)$ |
| --- | --- |
| $t_1 = \text{Aug. 7}$ | $n_1 = 11377 \pm 2205$ |
| $t_2 = \text{Aug. 14}$ | $n_2 = 18949 \pm 2652$ |
| $t_3 = \text{Aug. 21}$ | $n_3 = 27455 \pm 2281$ |

to yield a solution

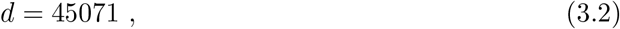

From Eq. (2.7) with *n* = 1 we get

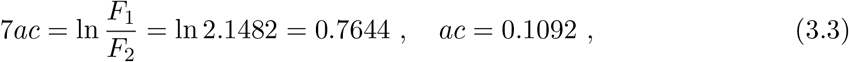

with *F*_1_ ≡ *F* (*t*_1_) and *F*_2_ ≡ *F* (*t*_2_).

Substituting the result *ac* = 0.1092 into Eq. (2.6), we have

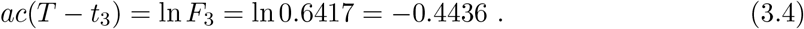

with *F*_3_ ≡ *F* (*t*_3_), to yield to yield *T* − *t*_3_ = −4.062, so that

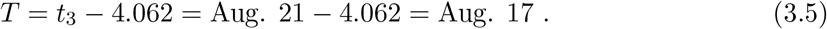

Error estimations for *d* and *T* can be seen from Appendix. By using relative errors.,

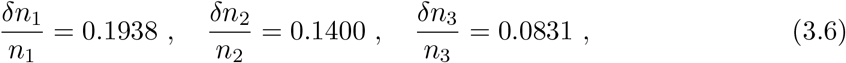

we have

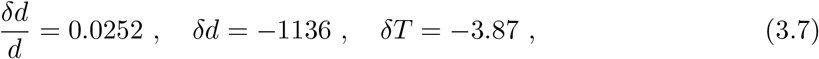

so that

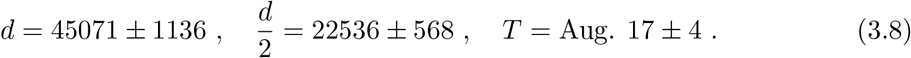

The value of *d* = 45071 on Aug.17 is plotted in Fig. 1. Values of *d* for the other *n* are also plotted in Fig. 1.

**Figure 1:**
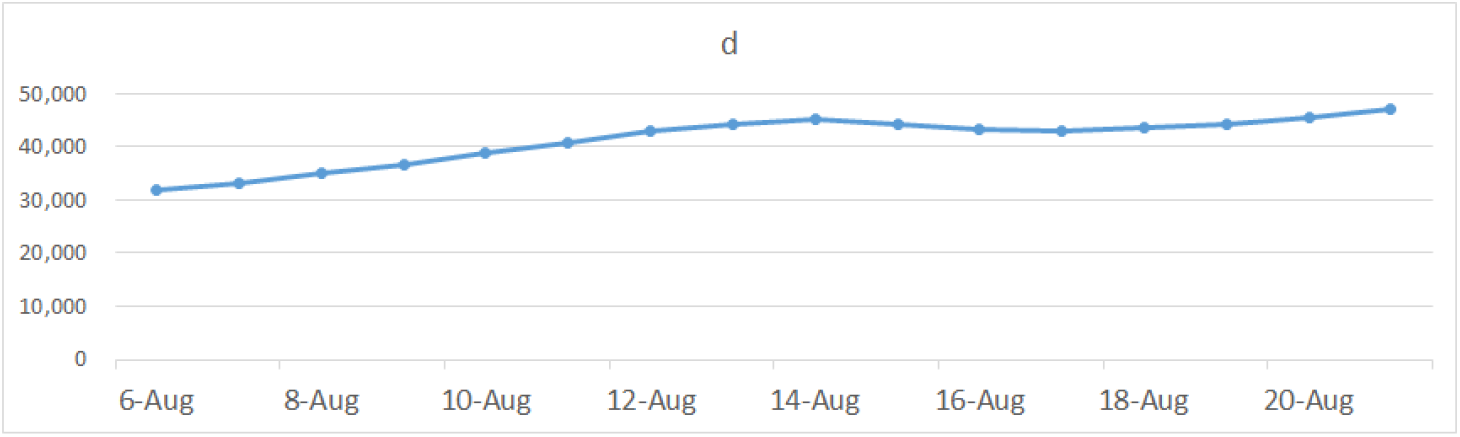
Blue points show values of *d*, which is defined by *d* = *R*(∞), e. g., the final total removed number, and is a solution of Eq. (2.10) for each *n*.

In Fig. 2 we draw a red curve of Eq. (2.5) for *R*(*t*) calculated with the average value *d* = 44465 and *ac* = 0.1106 from Aug.13 to Aug.20, while the blue curve shows data for *R*(*t*). Both lines coincide well in a region before Aug.27. The value of d should be constant as seen between Aug. 13 and Aug.20 within errors 1200~5000, but not before Aug.12. The deviation comes from the deviation between the red line and blue line in Fig. 2, where the red line is the calculated one of *R*(*t*) and the blue line is its data. From this reason we abandon data outside of Aug.13~20.

**Figure 2:**
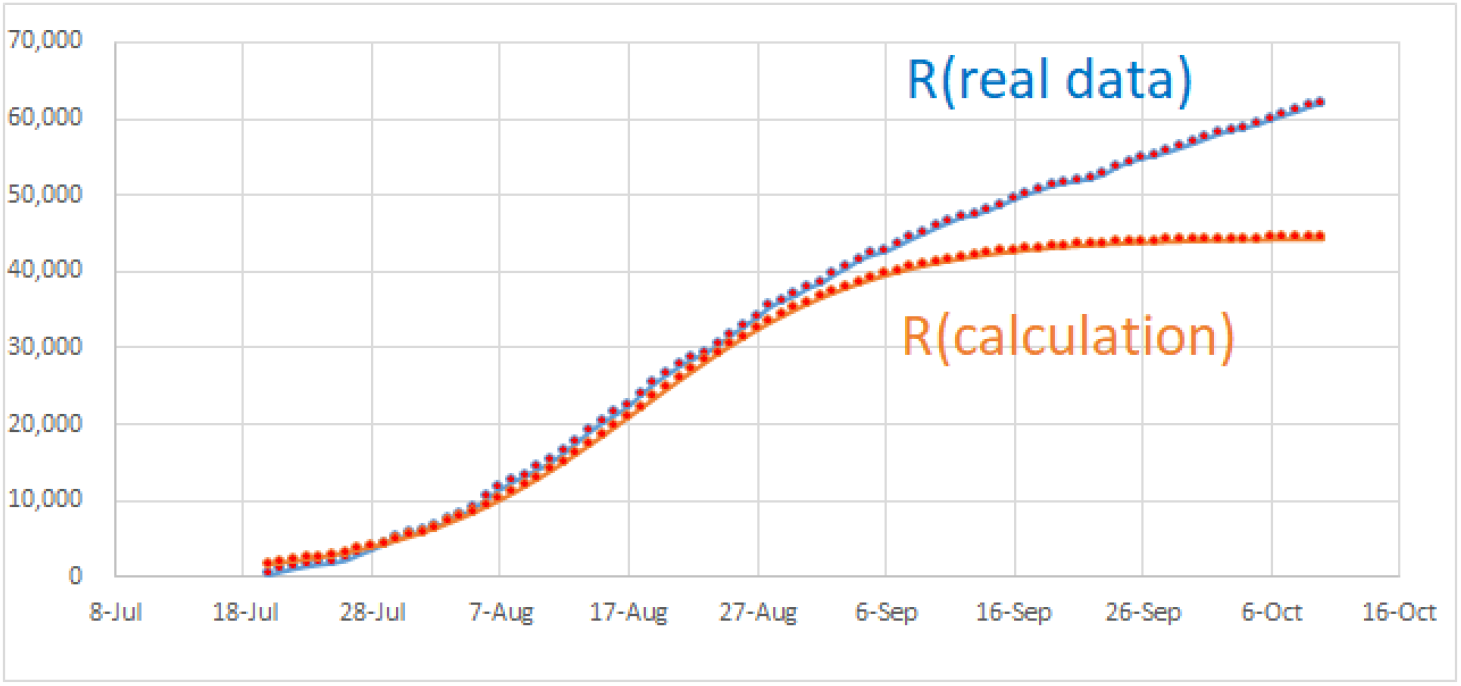
The red line is a calculated curve of *R*(*t*), while the blue line shows data for *R*(*t*). Both lines coincide well in a region before Aug. 27.

**Figure 3:**
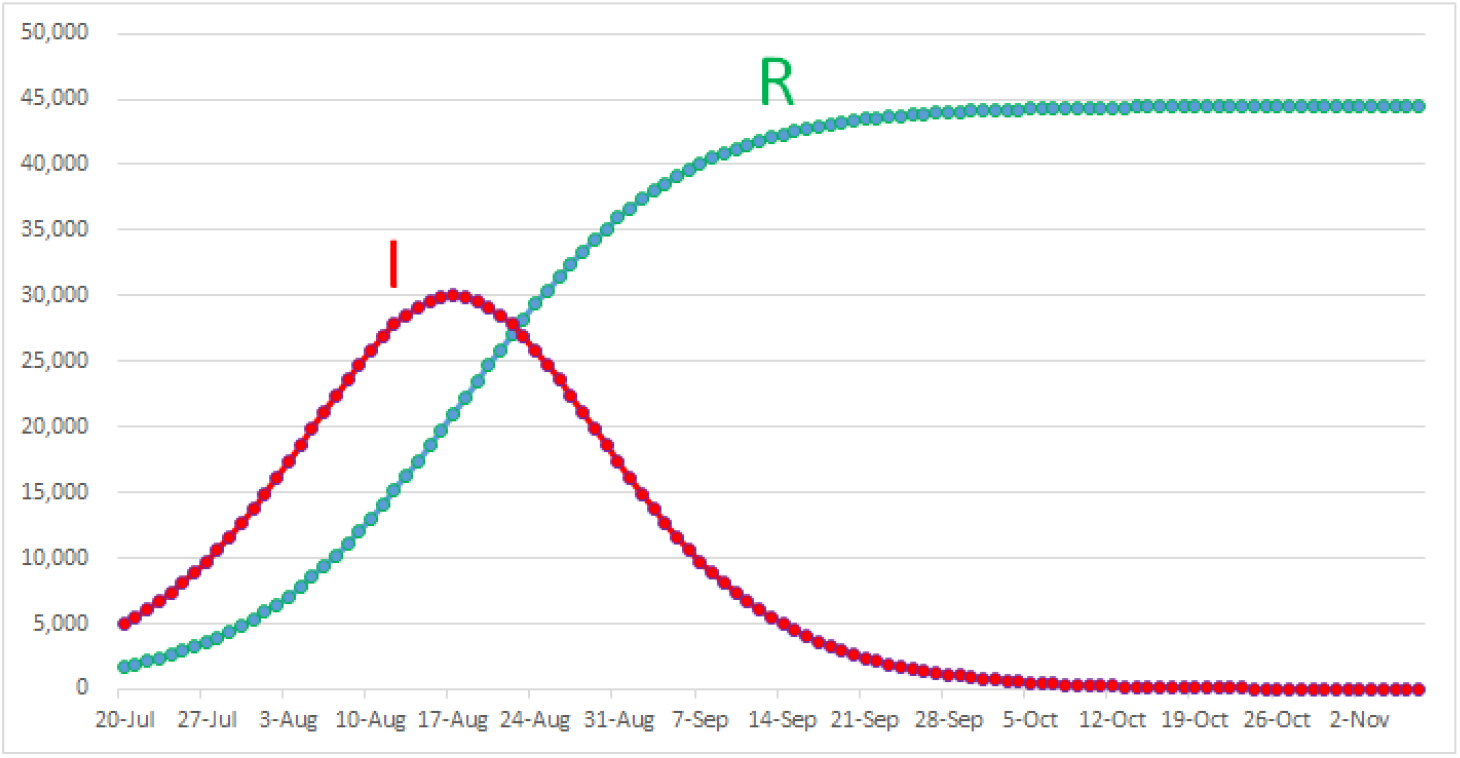
The blue line shows the calculated line of *R*(*t*). The red line is the infectious number *I*(*t*) with *d* = 4465 and *a* = 2.698.

To sum up, the second wave started from July.20, 2020, and peaked at Aug.17 with its total removed number 22536 ± 568. These calculated values should be compared with actual data that the peak date is around Aug.17 with its total removed number 22460.

## 4 Application to the third wave of COVID-19 in Japan

Our logistic formula is applied to the third wave of COVID-19 in Japan. This provides a revise of previous work [13]

The *R*(*t*) is the accumulated number of removed in the third wave in Japan, which is an average for 7 days in a middle at each t with standard deviations, where *t* is the date starting from Oct. 11. We have subtracted the accumulated number 82810 of removed in the first and second waves from that in the first, second and third waves.

Substituting data in the Table 2 into Eq. (2.10) with *n* = 1, we have the equation for *d*

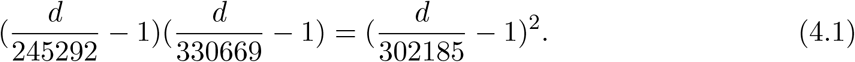

**Table 2:** Date and the removed number in the third wave in Japan [14]

| t | D(t) |
| --- | --- |
| $t_1=\text{Jan. 29}$ | $n_1 = 245292 \pm 11265$ |
| $t_2=\text{Feb. 12}$ | $n_2 = 302185 \pm 5537$ |
| $t_3=\text{Feb. 26}$ | $n_3 = 330669 \pm 3186$ |

to yield a solution

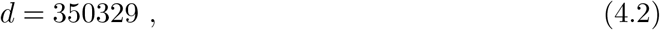

From Eq. (2.7) with *n* = 1 we get

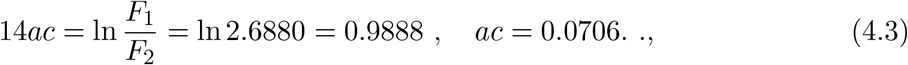

Substituting the result *ac* = 0.0701 into Eq. (2.6), we have

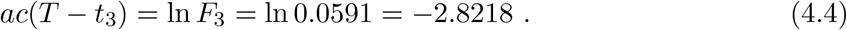

to yield to yield *T* − *t*_3_ = −39.97, so that

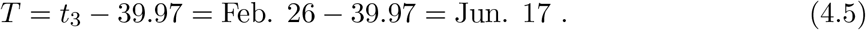

Error estimations for *d* and *T* can be seen from Appendix. By using relative errors.,

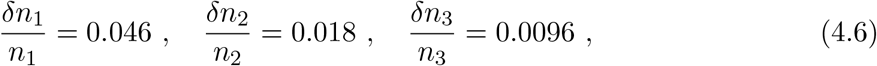

we have

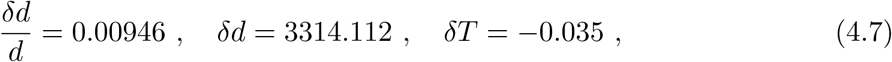

so that

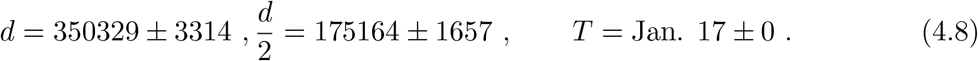

The value of *d* = 350329 on Feb. 12 is plotted in Fig. 4. Values of *d* for the other *n* are also plotted in Fig. 4.

**Figure 4:**
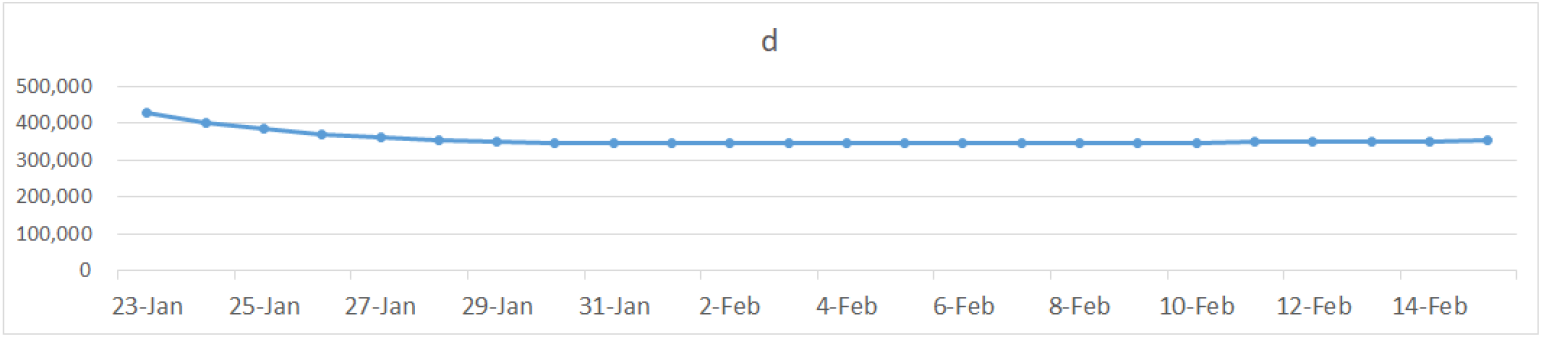
Blue points show values of *d*, which is defined by *d* = *R*(∞), e. g., the final total removed number, and is a solution of Eq. (2.10) for each *n*.

In Fig. 5 we draw a red curve of Eq. (2.6) for *R*(*t*) calculated with the average value *d* = 348008 and *ac* = 0.0723 from Jan. 28 to March 15, while the blue curve shows data for *R*(*t*). Both lines coincide well in a region after Jan. 19. The value of d should be constant as seen after Jan. 24, but not before Jan. 24. The deviation comes from the deviation between the red line and blue line before Jan. 24 in Fig. 5, where the red line is the calculated one of *R*(*t*) and the blue line is its data. From this reason we abandon data before Jan.24.

**Figure 5:**
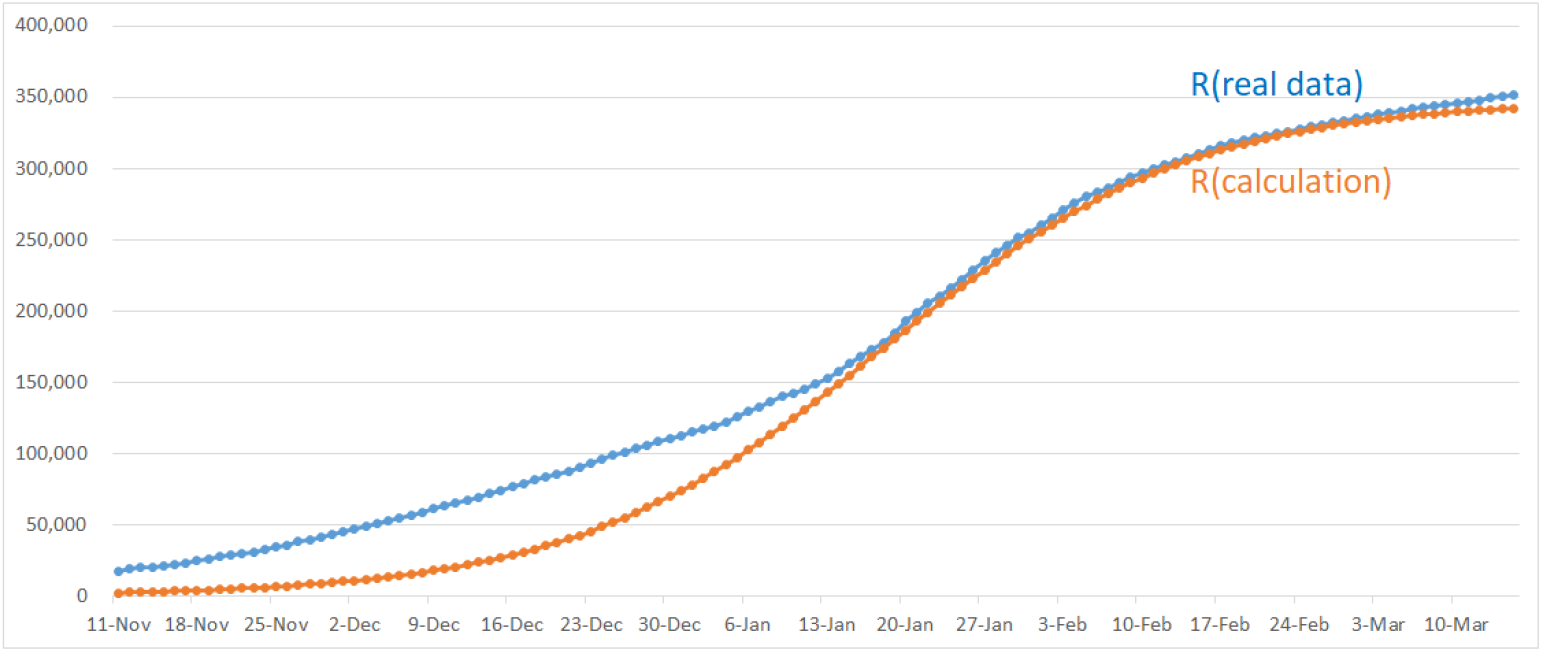
The red line is a calculated curve of *R*(*t*), while the blue line shows data for *R*(*t*). Both lines coincide well in a region after Jan. 24.

**Figure 6:**
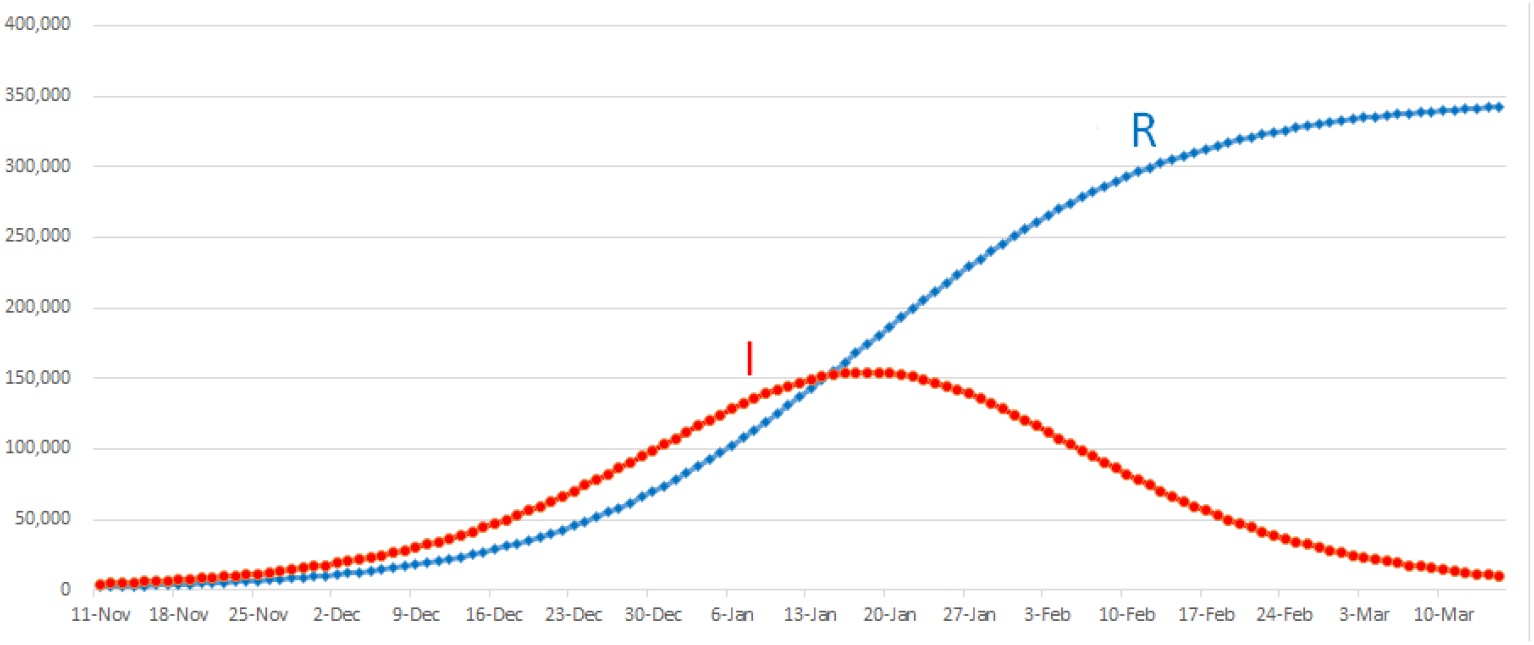
The blue line shows the calculated line of *R*(*t*). The red line is the infectious number *I*(*t*) with *d* = 348008 and *a* = 1.763.

To sum up, the third wave started from Oct.11, 2020, and peaked at Jan. 17 with its total removed number 175165 ±1657. These calculated values should be compared with actual data that the peak date is around Jan. 17 with its total removed number 177501.

## 5 Concluding remarks

The logistic formula in biology is applied, as the first principle, to analyze the removed number by the second and third waves of COVID-19 in Japan.

The second wave started from July.20, 2020, and peaked at Aug.17 with its total removed number 22536 ± 568. These calculated values should be compared with actual data that the peak date is around Aug.17 with its total removed number 22460.

The third wave started from Oct.11, 2020, and peaked at Jan. 17 with its total removed number 175165 ± 1657. These calculated values should be compared with actual data that the peak date is around Jan. 17 with its total removed number 177501. Results of the third wave have been obtained by using new data after the peak, Jan. 17. So, these are not a kind of prediction. However, we have succeeded to reproduce the peak data fairly well.

## Data Availability

the availability of all data is yes.

https://toyokeizai.net/sp/visual/tko/covid19

## Acknowledgement

We would like to thank K. Shigemoto for many valuable discussions and big supports.

## Appendix Error estimation formulas

From the formula 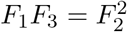 the relative error of removed number *d* is driven as

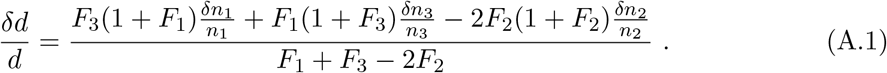

Secondly, the equation for the peak day *T, ac*(*T* − *t*_3_) = ln *F*_3_, yields a formula

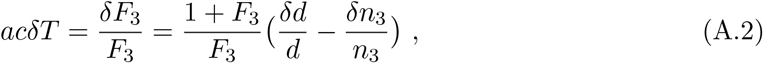

from which one can estimate *δT*.

